# The Impacts of Testing Cadence, Mode of Instruction, and Student Density on Fall 2020 COVID-19 Rates On Campus

**DOI:** 10.1101/2020.12.08.20244574

**Authors:** Christopher W. Stubbs, Michael Springer, Tasha S. Thomas

## Abstract

We analyzed the COVID-19 infection rate among undergraduate students at 9 colleges and Universities in the greater Boston area and 4 comparison schools elsewhere, from Fall 2020. We assessed whether the cumulative rate of infection is dependent on the mode of instruction (in-person, hybrid, or remote), on the number and density of dorm-resident undergraduates, and/or on COVID-19 testing cadence. We limited our analysis to institutions that have implemented at least weekly PCR testing of dormitory-resident undergraduates. Our primary conclusions are that (i) the fraction of students that succumbed to a COVID-19 infection up through Nov 22, 2020 shows no correlation with either the total number of students on campus, or the fractional occupancy of dormitories, (ii) remote instruction vs. hybrid instruction has no significant measurable impact on cumulative infection rate, and (iii) there is evidence that testing 2 or 3 times per week is correlated with lower infection rates than weekly testing. These data are consistent with a hypothesis of students predominantly acquiring infection off-campus, with little community transmission within dormitory housing. This implies good student compliance with face mask and social distancing protocols.

**Significance Statement:** We review the incidence of COVID-19 infection among under-graduate students for selected colleges and universities that conducted at least weekly COVID-19 testing during the Fall of 2020. We analyzed the infection-rate dependence on number of students on campus, dormitory residential density, instructional methodology (remote vs. hybrid), and testing cadence. This compilation of outcomes can help inform policy decisions for congregate settings.

---

**M**any colleges and universities are making plans for Spring 2021, and beyond. Faced with a rising number of COVID-19 infections nationwide, campus leaders are keen to understand how the mode of instruction and student residential policies impact the transmission of the virus within their undergraduate population. We have gathered publicly available data up through Nov 22, 2020 from nine institutions of higher education in the greater Boston area, and four from outside this region.

The transmission of COVID-19 in congregate housing settings is a source of considerable concern. Moreover, prior to the start of the 2020-2021 academic year there was apprehension (1) that large groups of students returning to the Boston area could trigger significant local COVID-19 outbreaks.

The availability of significant RT-PCR (2) testing capacity in the Boston area allowed regional institutions of higher education to undertake high-cadence COVID-19 surveillance campaigns for their students. The Broad Institute implemented (3) sufficient testing capacity to serve over 100 institutions, with the ability to process up over 100,000 tests/day. In addition, Boston University (BU) and Northeastern University implemented (4, 5) the capacity to process 6,000 tests/day to meet the needs of their community. The testing performed by the higher education community comprises a significant fraction of all the COVID-19 testing undertaken in the Commonwealth of Massachusetts.

To mitigate viral spread, high cadence PCR testing of campus-residential students is accompanied by prompt isolation of those individuals whose samples contain SARS-COV2 RNA. In addition, contact tracing identifies and quarantines people who are deemed close contacts, who are at increased risk of both infection and infecting others. Importantly, if done rapidly enough (6–8) these contacts can be sequestered before they enter the infectious period of the disease. This strategy is most effective (9) if the close contacts are quarantined in less time than the latency period (the time interval between being infected and becoming infectious), which for COVID-19 is thought to be a few days (10). These are among the considerations that determine the testing cadence choices made by institutions.

## 1. Data

Table 1 presents the information we compiled from institutional web sites and press releases. We determined the total number of undergraduate students (N_*s*_) in campus housing, the fractional occupancy (*ρ*) of dormitories, the cadence (T) of COVID-19 surveillance tests for the students, the cumulative number (N_*i*_) of COVID-19 infections detected in the students between mid-Aug and Nov 22, 2020. From these numbers we computed *f*, the fraction of on-campus students who tested positive for COVID-19 during the interval in question, expressed as infections per thousand students. The definition of an infection is a positive PCR COVID-19 test result, as reported by the institution.

**Table 1.** Input Data. The columns list (1) number of undergraduate students in residence, N_*s*_, rounded to the nearest hundred (2) dormitory fractional occupancy density *ρ* (3) COVID-19 test cadence (tests/student/week), T, (4) instructional mode, (5) cumulative number of undergraduate COVID-19 infections Aug 15, 2020 through Nov 15, 2020, N_*i*_, and (6) cumulative fraction of dorm-resident students (per thousand) that tested positive over that interval, *f*.

| Institution | $N_s$ | $\rho$ | $T$ | Mode | $N_i$ | $f$ |
| --- | --- | --- | --- | --- | --- | --- |
| **Boston Area** |  |  |  |  |  |  |
| Boston Coll. (11) | 9400 | 1.00 | 1 | Hybrid | 275 | 30 |
| Boston Univ. (12) | 16800 | 0.25 | 2 | Hybrid | 354 | 21 |
| Brandeis Univ. (13) | 1900 | 0.50 | 2 | Hybrid | 32 | 17 |
| Emerson Coll. (14) | 3800 | 1.00 | 1 | Hybrid | 32 | 8 |
| Harvard Coll. (15) | 1500 | 0.25 | 3 | Remote | 26 | 17 |
| Northeastern Univ. (16) | 19900 | 1.00 | 2 | Hybrid | 274 | 14 |
| Tufts Univ. (17) | 6000 | 1.00 | 2 | Hybrid | 98 | 16 |
| U. Mass Boston (18) | 1000 | 1.00 | 1 | Hybrid | 26 | 25 |
| Wellesley Univ. (19) | 2400 | 1.00 | 2 | Hybrid | 7 | 3 |
| **Non-Boston** |  |  |  |  |  |  |
| Columbia Univ. (20) | 900 | 0.15 | 1 | Remote | 27 | 30 |
| Cornell Univ. (21) | 5100 | 0.73 | 2 | Remote | 175 | 34 |
| Amherst Coll. (22) | 1000 | 0.60 | 3 | Hybrid | 4 | 4 |
| Williams Coll. (23) | 1500 | 0.75 | 2 | Hybrid | 7 | 5 |

We drew upon publicly available data for the period Aug 15 through Nov 22, 2020. Most institutions sent students home at the start of the 2020 Thanksgiving weekend, so this spans the time most undergraduates were in residence for Fall 2020.

In order to factor out variability in the underlying local rate, we selected a variety of Boston-area institutions^∗^ as well as a few large and small institutions in other urban and rural areas. Since we are particularly interested in investigating the possible impact of testing cadence, we limited our selection to schools that have at least once per week PCR tests (unpooled) for their residential undergraduates.

The numbers in Table 1 include some COVID-19 cases detected upon student arrival (that students brought to campus with them) and are therefore an upper bound on transmission on-campus.

## 2. Analysis

Our goal was to explore how the rate of infections acquired by campus-resident undergraduate students depended on (i) the number and density of students living on campus, (ii) the method of instruction for Fall 2020, and (iii) the frequency of COVID-19 surveillance testing. There are numerous other important and potentially confounding factors that we have not taken into account, such as ventilation, mask compliance, dining policy, symptom attestation, turnaround time, isolation/quarantine latencies, students per bedroom, sample collection methodology and effectiveness, and the social network structure of the interactions within the student population, *e*.*g*. whether students were grouped into “pods.”

Small-number statistics can obscure trends; a single “superspreader” event can distort the picture. Nevertheless we think the exploration presented here is informative.

We focused on Boston-area institutions to compensate for variations in the underlying rates of COVID-19 infection in the surrounding community. Aggregating statistics on a national scale could run the risk of the institution-policy-dependence being obscured by variations in the surrounding regional circumstances.

If all the acquired infections were statistically independent, *i*.*e*. were the result of off-campus interactions in the community, the fractional incidence in the student groups should be independent of both total population size and dormitory density. If, on the other hand, one were to see a strong dependence on dormitory density, that might support a hypothesis of transmission due to congregate housing and proximity.

The interval in question spans 100 days. An estimate of the COVID-19 prevalence in the Boston-area residential undergraduates, expressed as new cases per day per 100,000 people, is then our normalized *f*, the cumulative number of infections per thousand students over the 100 day period. The mean of *f* for the Boston area schools is *< f >*= 16*±*3 new cases per 100,000 person-days. The uncertainty was computed by taking the standard deviation of the Boston-area *f* values and dividing by 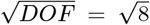, where *DOF* is remaining degrees of freedom, This is consistent with the mean case rate of 10.8 per 100,000 reported (24) for Middlesex County, MA over the same period. The local colleges contribute a very small fraction of the countywide case totals, and don’t distort that statistic. Since the on-campus testing campaigns would detect asymptomatic cases, who are presumably under-represented in the regional case rate estimates, one might expect the case rate determined from campus surveillance to exceed the local regional rate, due to higher completeness. Making a correction for selection effects in the regional case rate requires better data than is currently available.

### A. Dependence on Total Student Population and Dormitory Occupancy Density

Figure 1 shows the fraction *f* of dorm-resident students who tested positive over the study interval vs. total number of students in residence on each campus. Figure 2 shows how *f* depends on dormitory occupancy density.

**Fig. 1.**
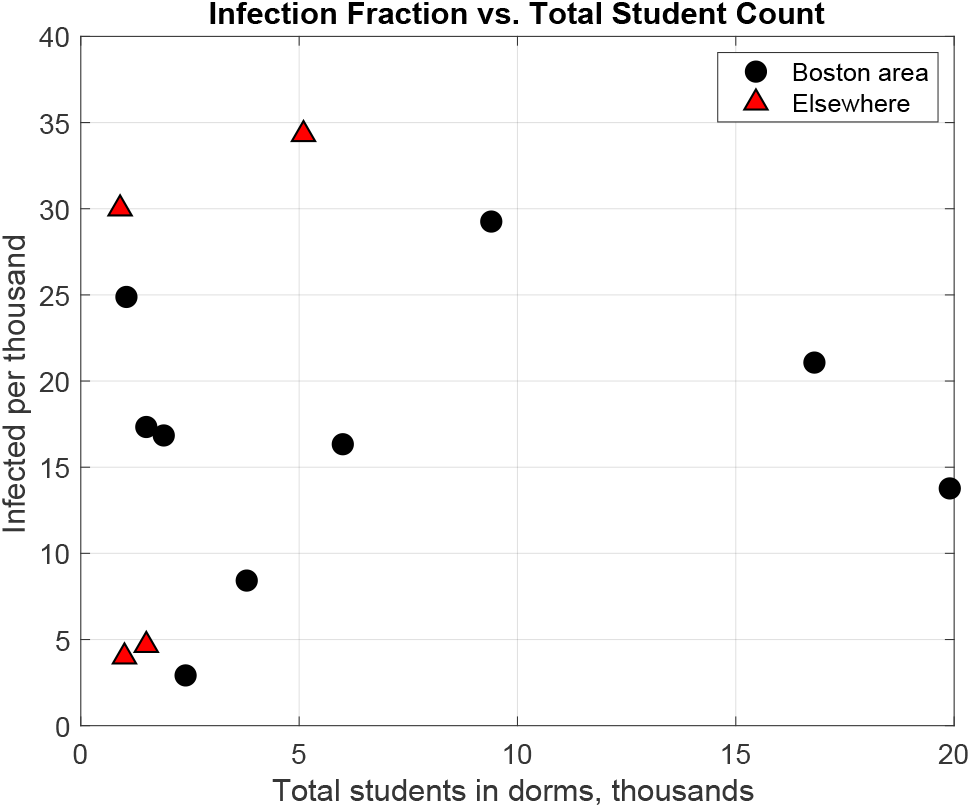
Cumulative infected student fraction *f* vs. total students in residence. The vertical axis shows the total reported infections per thousand students. The horizontal axis is the total student dorm-residential population. Boston area schools are shown as circles, and comparison schools from other areas as triangles.

**Fig. 2.**
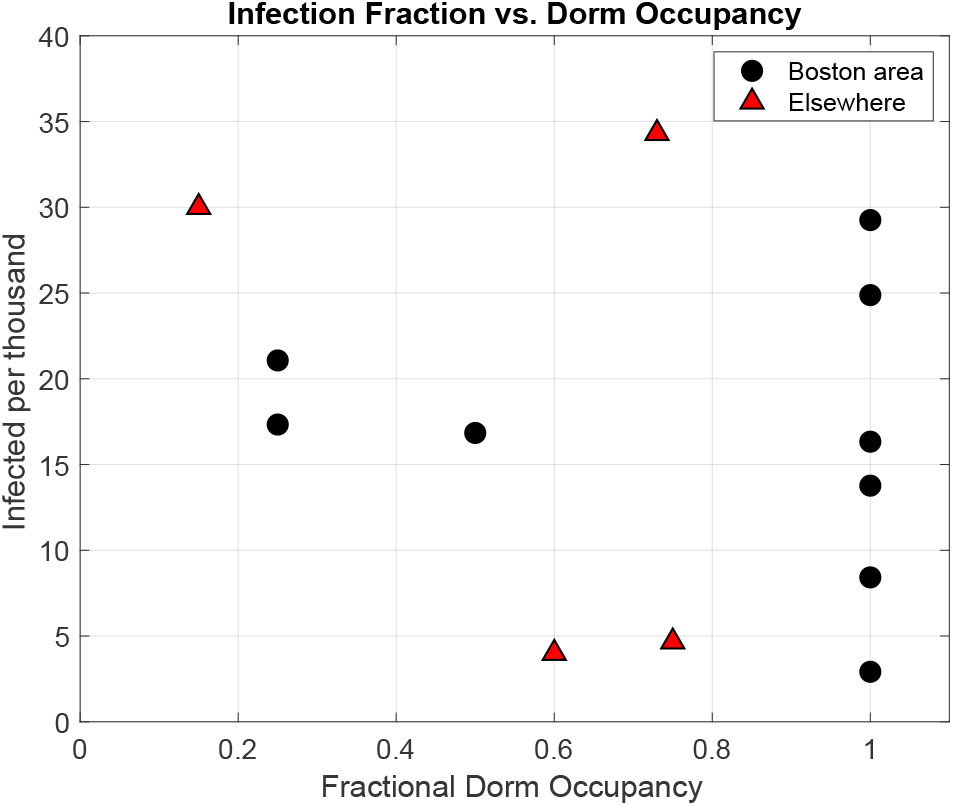
Cumulative infected fraction *f* vs. dormitory density *ρ*. The vertical axis shows the total reported infection per thousand students. The horizontal axis is the dormitory residential density. Boston area schools are shown as circles, and comparison schools from other areas ass triangles.

There is no apparent correlation of *f* with either the number of students in residence or the fractional occupancy density of dormitories. The correlation values of *r*^2^ are shown in Table 2. The scatter in the data appears to be driven by factors other than total students on campus and the density of dormitory occupancy.

**Table 2.** Correlation of cumulative infection fraction *f* vs. number of students in residence, and against dormitory occupancy density. These are the relationships plotted in Figure 1 and 2. There is no apparent correlation, for either the Boston-area subset or the full sample. The columns show the subsample used, the values of the correlation coefficient r^2^, and the number of degrees of freedom DOF for each comparison.

| Correlation | $r^2$ | DOF |
| --- | --- | --- |
| Boston area, total students | 0.03 | 7 |
| Boston area, dorm fraction occ. | 0.03 | 7 |
| Full sample, total students | 0.02 | 11 |
| Full sample, dorm fraction occ. | 0.06 | 11 |

### B. Dependence on Mode of Instruction

We explored the extent to which mode of instruction is correlated with infection rates. “Remote” means no in-person interactions, with all classes conducted online. “Hybrid” instruction includes some element of in-person classroom interaction. Figure 3 shows the dependence of *f* vs. mode of instruction.

**Fig. 3.**
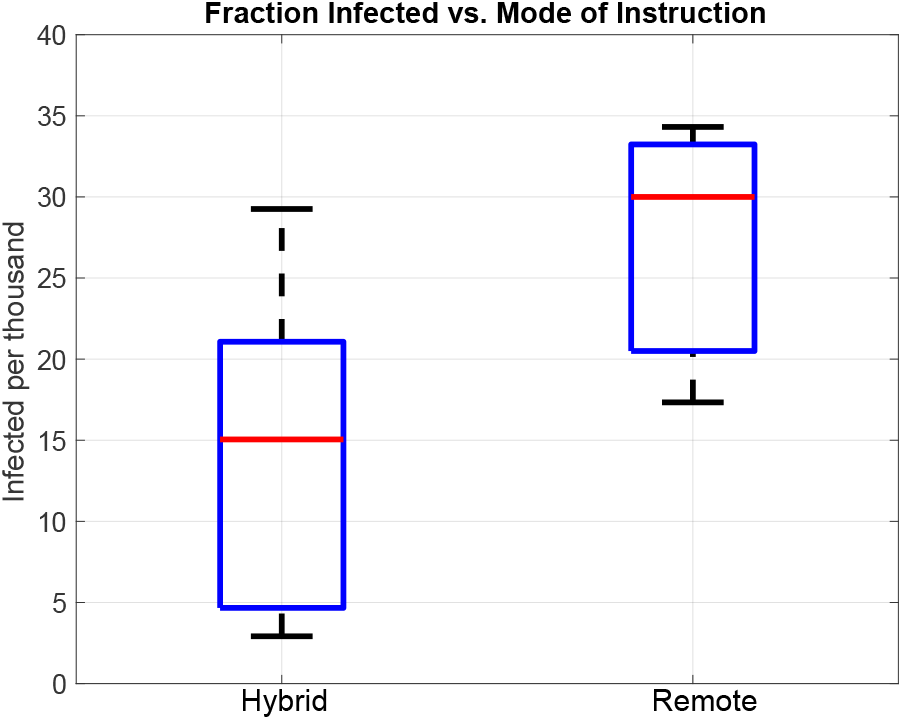
Box plot of cumulative normalized infection *f* as a function of style of instruction. Remote-only is shown on the right, with mixed or hybrid instruction on the left. None of the institutions studied had in-person-only instruction. The red bars show the median values, the blue boxes span the 25^th^ to 75^th^ percentiles, and the grey bars span the full range of the respective distributions. A t-test indicates no statistically significant difference between the means as a function of mode of instruction.

A two-sample t test (MATLAB t test2, allowing for unequal variances) against mode of instruction returns a 5.3% likelihood of the two means being the same, *i*.*e*. there is at best weak evidence of an infection rate difference that is correlated with the mode of instruction. This test was carried out on the full sample. Since the sense of the correlation indicates Remote instruction correlates with higher infection, there is no evidence that Hybrid instruction leads to increased COVID-19 cases.

### C. Dependence on Testing Cadence

Figure 4 shows the dependence of cumulative infection fraction *f* vs. viral test cadence.

**Fig. 4.**
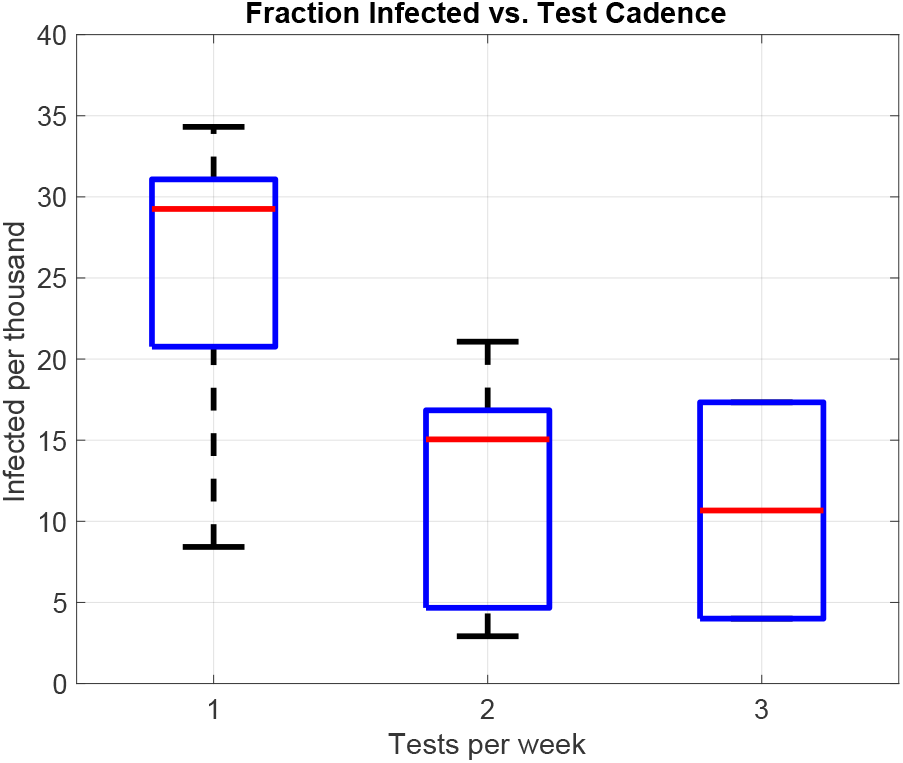
Box plot of cumulative relative infection *f* as a function of PCR test cadence. There is evidence (p-value of 1.7%) that a campus testing frequency of more than once per week is correlated with lower *f*. There were only two institutions in our sample that carried out testing three times per week, so the horizontal bars of the blue box on the right correspond to those values, and the red bar is their mean.

Since only two of the schools in our sample carried out testing three times per week, we binned the cadences into once per week, and more than once per week. A two-sample t test against this cadence parameter returns a p-value of 1.7%, which does imply a correlation where schools that tested 2 or 3 times per week had fewer infections per capita than schools that conducted weekly tests. This comparison was carried out on the full sample.

## 3. Conclusions

While the statistical power of this analysis is limited, the results are consistent with the hypothesis that most student infections were acquired outside of the dorm-residential setting, with minimal community transmission within congregate on-campus student housing. This is consistent with anecdotal contact tracing reporting on our campus.

While there could have been a strong correlation of infection rate with the housing variables we considered, this turned out to not be so. The number of serologically-confirmed COVID-19 infections that manifest in the students that were sequestered into quarantine due to close contacts will help determine the extent to which “test+isolate+quarantine” (TIQ) strategies suppressed subsequent transmission. That analysis must also take into account the false-positive rate from PCR testing; the frequency of this is presently unclear.

Great effort should be made to minimize the latencies in the TIQ timeline. This timeline starts when the students collects the sample, and ends when infected students are in isolation and their close contact have been placed in quarantine. Our experience indicates that estimates of these latencies in earlier modeling may have been over-optimistic.

We see a correlation between testing more frequently than once per week and lower COVID-19 rates in campus-resident undergraduates. This result is consistent with earlier modeling (*e*.*g. (25)*). This correlation is insufficient to conclude that the higher testing cadence alone directly suppressed COVID-19 transmission. Schools that did testing more often may also have implemented more restrictive social policies, and made *e*.*g*. ventilation upgrades that led to lower rates of infection.

The Boston area institutions of higher education that implemented at least weekly viral testing appear to have avoided both (i) an incidence of COVID-19 infections that substantially exceeded that of the surrounding community, and (ii) any sustained uncontrolled outbreak in on-campus congregate housing.

The relationship between institutional policies and outcomes warrants further study. Future analyses will benefit from ready access to the relevant data. We advocate an effort to provide open access to standardize data structures that would include:

- student housing: number of students, housing density, numbers per bedroom, numbers per bathroom, etc.
- face mask policies, and compliance estimates,
- daily testing record and associated attributes (numbers tested from on-campus and off-campus residential populations, undergraduate, graduate, postdocs, faculty, staff, and professional student status, *etc*.),
- daily log of residential and non-residential students in isolation and quarantine, and
- mode of instruction: distributions of number of students engaged in in-person, hybrid, and fully remote learning.
- metrics that capture the social limitations put in place by the institution, and compliance indicators.
- residential ventilation characteristics, such as cubic feet per minute of HEPA-quality filtered air exchange.

It is presently unclear how rapidly campus-resident populations will be vaccinated against COVID-19. An ongoing assessment of infection rates as a function of institutional policy will help inform evidence-based decisions and the agile implementation of best practices.

## Data Availability

Data was downloaded from the cubically available dashboard for universities. Code available upon request.

## ACKNOWLEDGMENTS

We gratefully acknowledge the students whose conduct and adherence to public health protocols during Fall 2020 helped achieve the results presented here. We thank E. Carrington Gregory for suggestions on the manuscript. We also thank Harvard College Dean of Students, Katherine G. O’Dair for helping us to compile additional university data from Columbia College and Cornell University. Our work was supported by Harvard University.

## Footnotes

∗ We were unable to determine the number of positive results for residential students at MIT, so it’s not included here.

